# Trends and epidemiological profile of preventable hospitalizations in Honduras (2014 – 2024): An 11-year analysis of ambulatory care sensitive conditions

**DOI:** 10.64898/2026.04.22.26351522

**Authors:** Héctor Enrique Alfaro, Jonathan Lara-Arevalo

**Author notes:** **Corresponding author:** (JLA).

## Abstract

Ambulatory Care Sensitive Conditions (ACSCs) are conditions for which effective and timely primary health care (PHC) can prevent hospitalizations. They are widely used as a proxy indicator of access to and quality of PHC. Despite their relevance, evidence from Central America remains scarce. This study aimed to quantify the burden, describe the epidemiological profile, and assess temporal trends of ACSCs hospitalizations in Honduras from 2014 to 2024. We conducted a retrospective observational study using national administrative hospital discharge data from all Ministry of Health hospitals. ACSCs were defined using a standardized list of 20 diagnostic groups based on ICD-10 codes. We estimated percentages and sex-age-standardized hospitalization rates per 10,000 inhabitants. Clinical indicators included length of stay (LOS) and in-hospital fatality rates. Temporal trends were evaluated using joinpoint regression models to estimate annual percent changes (APC). Analyses included stratification by age, sex, and disease category. A total of 4,023,944 hospitalizations were analyzed, of which 547,486 (13.6%) were classified as ACSCs. The overall sex-age-standardized rate was 54.1 per 10,000 inhabitants. ACSCs’ standardized rates increased between 2014 and 2018 (APC: 2.7%; 95% CI: -2.4; 15.2), declined sharply between 2018 and 2021 (APC: −17.8%; 95% CI: −30.6; −10.3), and increased again between 2021 and 2024 (APC: 15.9%; 95% CI: 4.6; 37.6). Despite this rebound, rates remained below pre-pandemic levels. ACSCs were concentrated among children under 5 years (27.7%) and adults aged 60 years and older (29.9%). Noncommunicable diseases accounted for 56.8% of cases, with diabetes mellitus as the leading cause. Compared with non-ACSCs hospitalizations, ACSCs were associated with longer LOS (4.9 vs. 3.9 days; p <0.001) and higher in-hospital fatality rates (2.4% vs. 1.7%; p <0.001). ACSCs hospitalizations constitute a substantial burden in Honduras and reflect persistent gaps in PHC performance. Strengthening PHC resilience and capacity, particularly for chronic disease management and vulnerable populations, is essential to reduce avoidable hospitalizations and improve health system efficiency and equity.

## Introduction

In Latin America and the Caribbean (LAC), progress toward Universal Health Coverage has coexisted with fragmented health systems and persistent gaps in access to and quality of care. [1] In parallel, out-of-pocket expenditure remains high (accounting for approximately 30.0% of total health expenditure in 2022), reflecting persistent economic barriers to timely access and limitations in ensuring financial protection. [2] These challenges demonstrate the need to strengthen health system organization and improve the capacity of primary care services to deliver timely and effective care [3].

Primary Health Care (PHC) is recognized as the strategy for organizing Integrated Health Service Networks, improving outcomes, and reducing inequities. [4] International and regional evidence has consistently documented that systems with strong PHC are associated with lower mortality, greater equity, and improved efficiency, largely through enhanced care coordination and continuity. [5] In the region, strengthening PHC is a necessary condition for advancing toward integrated services, preventing avoidable hospitalizations for conditions manageable in ambulatory settings, and consolidating progress toward Universal Health Coverage [4–6].

In this context, the analysis of preventable hospitalizations for Ambulatory Care Sensitive Conditions (ACSCs) has consolidated as a widely used indicator to indirectly evaluate the performance and resolution capacity of the first level of care. Systematic monitoring of ACSCs hospitalizations allows for the identification of gaps in access to and quality of ambulatory care and can inform targeted interventions to strengthen the PHC’s resolution capacity and improve health system efficiency. [7–10] The underlying logic is that timely, continuous, and high-quality care at the primary care level can prevent unnecessary hospital admissions for various health conditions. [7,11]

The scope of results from this type of analysis has enabled this methodology to gain international recognition as a public policy tool for monitoring health system performance and comparing progress across countries. [7,8] For example, the Pan American Health Organization (PAHO), in its 2020–2025 Strategic Plan, highlighted the importance of monitoring ACSCs hospitalizations and set a target of a 10% reduction. [12] To achieve this, PAHO proposed a standardized set of 20 diagnostic groups based on the International Statistical Classification of Diseases and Related Health Problems, 10th Revision (ICD-10) [6].

However, despite the growing international use of this indicator, evidence from Central America remains scarce, evidencing a significant knowledge gap. Most ACSCs studies have been conducted in high-income countries or selected LAC settings, while few analyses in Central America have used nationally representative administrative data. Published evidence is limited to El Salvador and Costa Rica [6,13].

In Honduras, the lack of up-to-date, nationally representative studies limits the system’s ability to monitor trends in ACSCs hospitalizations and to design targeted policy strategies. In this sense, the availability of a national administrative database capturing all hospital discharges from the Ministry of Health (MoH) network offers a unique opportunity to address this gap. Additionally, the COVID-19 pandemic may have modified patterns of healthcare utilization and access to PHC services, making it relevant to analyze trends before, during, and after the pandemic period [1].

Therefore, this study aimed to quantify the magnitude, describe the epidemiological profile, and evaluate temporal trends of ACSCs hospitalizations in Honduras between 2014 and 2024. Specifically, we estimated the percentage and population-adjusted rates of ACSCs hospitalizations, described their epidemiological profile according to demographic and clinical characteristics, and examined variations across pre-pandemic, pandemic, and post-pandemic periods. By using national MoH administrative data, this study provides the first comprehensive assessment of ACSCs hospitalizations in Honduras and generates policy-relevant evidence to inform efforts to strengthen primary care and improve health system performance.

## Methods

### Study design and context

We conducted an observational, retrospective study using hospitalization-level administrative data from the discharges reported by all 33 public hospitals in the subsystem managed by the Honduras MoH. The study analyzed hospitalizations from January 1, 2014, to December 31, 2024, spanning an 11-year period.

Honduras is a lower-middle-income country in Central America with a population that grew from approximately 8.6 million in 2014 to 10.7 million by 2024. [14] The health system is highly fragmented, consisting of a public sector led by the MoH and the Honduran Social Security Institute, and a smaller private sector. The MoH serves as the primary provider covering approximately 60% of the population, specifically targeting the uninsured and those living in vulnerable conditions. [15] Honduras’s health financing system is shaped by severe public underfunding, with public health investment reaching only 3.5% of the Gross Domestic Product. Consequently, out-of-pocket expenditure has become the dominant source of financing (51.1%) [16].

### Data source and study participants

The data used in this study came from the National Hospital Discharge Database by Primary Condition, managed by the Health Statistics Department of the Honduras MoH. This database contains hospital discharge records and includes information on patient demographic characteristics, primary discharge diagnosis, length of hospital stay (LOS), discharge condition (improved, cured, deceased, or same condition), and hospital facility characteristics (secondary or tertiary level of complexity). The database provided to the research team (accessed on November 5, 2025) was completely anonymized and contained no personal identifiers; therefore, the authors did not have access to information that could identify individual participants at any point during or after data collection. Consequently, the unit of analysis was the hospitalization episode, and it was not possible to track individual patients across multiple hospitalizations or distinguish between initial admissions and readmissions.

The study population consisted of 4,151,121 hospital care records, corresponding to 84.7% of all discharges reported in the country during the study period by the MoH subsystems and the Honduran Social Security Institute’s hospital network. [17] The data represented the universe of discharges within the analyzed health subsystem. The researchers included any patient with at least one day of stay in a public hospital and excluded those with LOS longer than 365 days.

During data cleaning, 3.1% of the records (127,178) were excluded. From them, 123,527 records corresponded to: a) those provided at the primary care level, b) those provided by observation services or other episodes that did not require a hospital bed, and c) records with LOS exceeding 365 days. Additionally, 3,651 records were excluded as they had missing values in key variables needed for analysis, such as coded diagnosis or sex. A complete-case approach was used for the analysis, given the low percentage of missing data (<0.1%).

### Definitions and classification

We defined ACSCs according to the classification proposed by Alfradique et al., [11] which is also used in the PAHO 2020-2025 strategic plan. [12] The operational definition used in this study included 20 groups of conditions, identified by 143 specific ICD-10 codes. The complete list of diagnostic groups and ICD-10 codes used to define ACSCs is presented in **S1 Table**. Based on this classification, hospitalizations were categorized into two mutually exclusive groups: hospitalizations with a primary diagnosis corresponding to an ACSC and hospitalizations due to other causes (non-ACSCs).

Furthermore, we organized the ICD-10 codes into three broad categories of diseases, following Bernal et al.’s proposal: preventable infectious diseases, noncommunicable diseases (NCDs), and maternal, child, and nutritional conditions (MCNC). [6]

### Statistical analysis

Descriptive analyses were conducted to characterize ACSCs hospitalizations over time. We calculated frequencies and percentages for categorical variables, while continuous variables were summarized using means and standard deviations. ACSC characteristics were described by age, age groups, sex, diagnostic groups associated with ACSCs, the three broad disease categories, LOS, in-hospital fatality rates, and hospital complexity level. We described data across three epidemiological periods: pre-pandemic (2014–2019), COVID-19 pandemic (2020–2021), and post-pandemic (2022–2024).

Annual hospitalization rates per 10,000 inhabitants were calculated using national population estimates from 2014-2024 obtained from the National Institute of Statistics of Honduras. [14] To ensure comparability, sex-age-standardized rates were calculated through the direct method, employing the 2019 Honduran population at the standard reference. For all standardization procedures, age was categorized into 5-year age groups. Population denominators used for rate calculations are shown in **S2 Table**.

We used the chi-square test to compare in-hospital fatality rates between ACSCs and non-ACSCs hospitalizations. The distribution of LOS (a continuous variable) between the ACSCs and non-ACSCs groups was compared using the non-parametric Wilcoxon rank-sum test (Mann-Whitney U test). A p-value < 0.05 was considered statistically significant.

To formally assess temporal trends in age-standardized ACSC hospitalization rates, empirical log-linear joinpoint regression models were utilized. The year was treated as a continuous independent variable. Trends were quantified using Annual Percent Changes (APC), the Average Annual Percent Change (AAPC) for the entire study period, and their respective 95% confidence intervals (CI). [18] Model selection and the identification of optimal joinpoints were performed using the Weighted Bayesian Information Criterion (Weighted BIC), allowing up to three joinpoints (four temporal segments). To accurately capture acute systemic disruptions caused by the COVID-19 pandemic, grid-search parameters were configured to ensure at least one observation between consecutive joinpoints. Stratified analyses, modeled as independent cohorts, were conducted by sex and disease category to identify trend heterogeneity and assess the differential impact of the pandemic across subpopulations.

Data consolidation was performed using structured templates in Microsoft Excel 365 for initial cleaning and verification stages. Data processing was conducted using RStudio statistical software (version 4.5.2). [19] The temporal trend analysis was executed using the Joinpoint Regression Program (version 6.0.1). [20] The analytical code can be shared upon communication with the study’s corresponding author.

### Ethical considerations

This study relied exclusively on the secondary analysis of administrative data obtained from the Honduran MoH. The dataset was fully anonymized prior to access, and the authors did not have access to information that could identify individual participants at any point. The research protocol was reviewed by the Research Ethics Committee of the Central American Technological University (UNITEC; IRB No. 00012967), which determined that the study does not constitute human subjects research and, therefore, does not require institutional review board approval or informed consent. This determination is documented in Minutes No. 002-2026 (January 15, 2026).

## Results

Between 2014 and 2024, a total of 4,023,944 hospitalizations were recorded in the MoH hospital network, of which 547,486 corresponded to ACSC hospitalizations, representing 13.6% of all admissions (**Table 1**). Compared with non-ACSC hospitalizations, ACSC cases involved older patients on average (mean age: 36.1 vs. 25.7 years) and were more frequently managed in secondary-level hospitals (72.4% vs. 61.0%). Across the full study period, ACSC hospitalizations accounted for 2,667,137 bed-days, representing 16.4% of total bed utilization. They were also associated with a greater clinical burden, including a longer LOS (mean: 4.9 vs. 3.9 days; p < 0.001) and a higher in-hospital fatality rate (2.4% vs. 1.7%; p < 0.001) (**Table 1**).

**Table 1.**
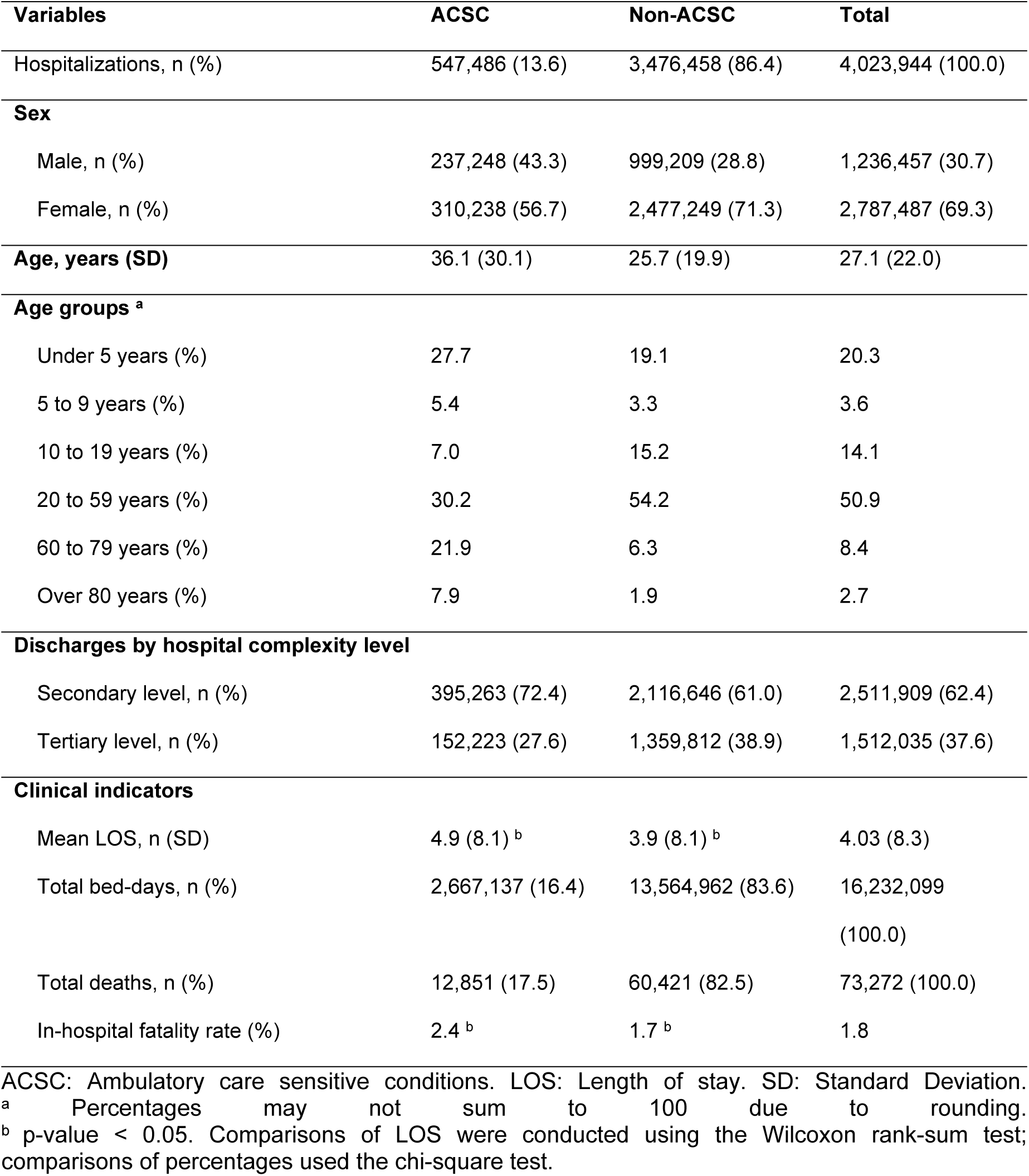
Comparative profile of ACSC vs. non-ACSC hospitalizations in Honduras, 2014–2024.

The overall sex-age-standardized ACSC hospitalization rate was 54.1 per 10,000 inhabitants, with notable variation across epidemiological periods (**Table 2**). In the pre-pandemic period, ACSCs accounted for 14.0% of hospitalizations, with a standardized rate of 62.8 per 10,000 inhabitants. During the COVID-19 pandemic (2020–2021), both the proportion and rate declined substantially to 10.8% and 35.6 per 10,000, respectively, before partially rebounding in the post-pandemic period to 14.3% and 50.7 per 10,000. Patterns in healthcare utilization were also reflected in hospital resource use and outcomes. The share of total bed-days attributable to ACSCs decreased during the pandemic and returned to pre-pandemic levels thereafter. Mean LOS for ACSC admissions remained relatively stable across periods, while in-hospital fatality rates increased from 2.1% in the pre-pandemic period to 2.8% in the post-pandemic period. Similarly, the ACSC to non-ACSC ratio declined during the pandemic and subsequently rebounded.

**Table 2.**
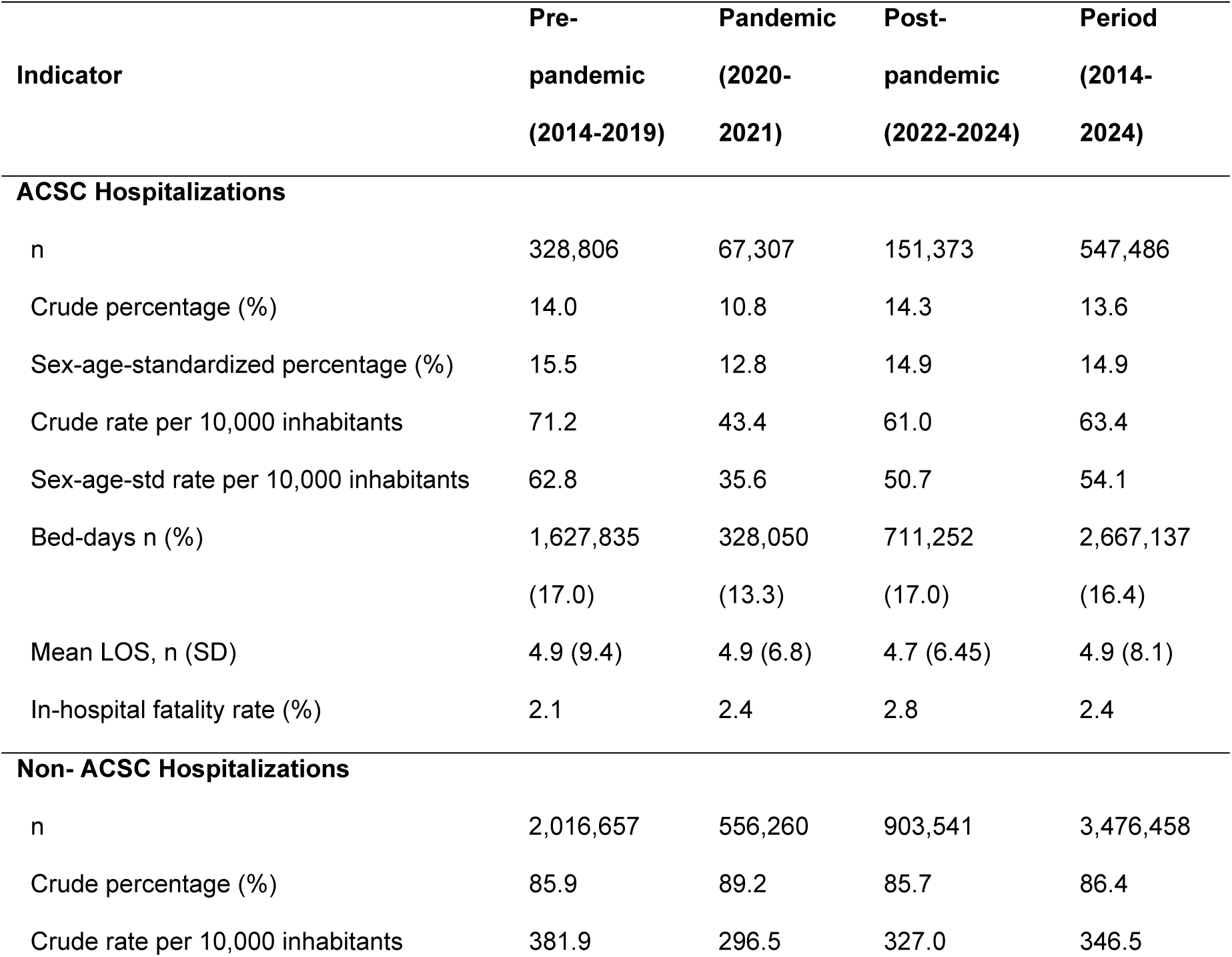
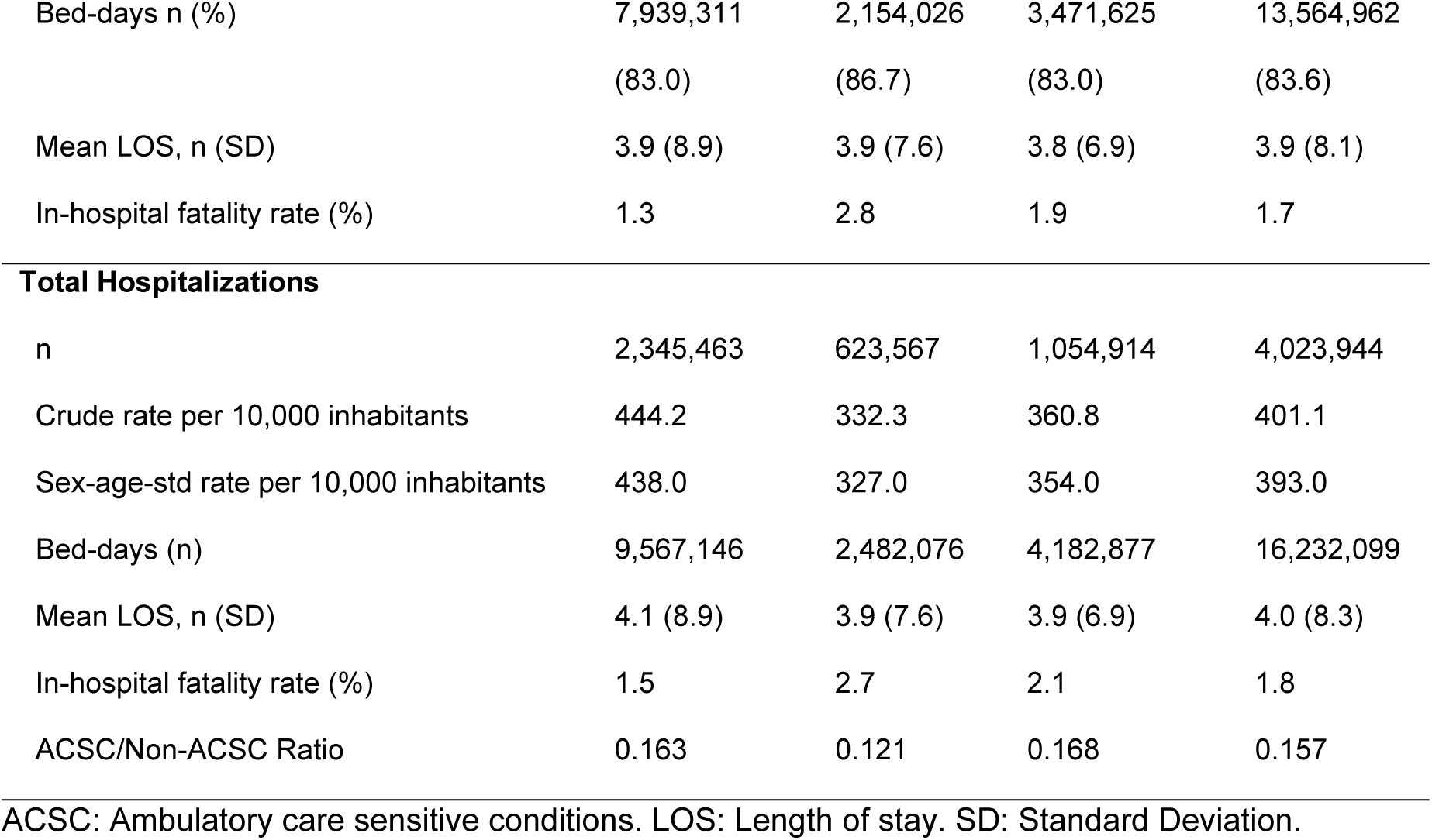
Hospitalization burden and ACSCs magnitude by epidemiological period (Pre-pandemic, Pandemic, and Post-pandemic), 2014–2024.

When categorized by disease group, NCDs emerged as the primary driver of ACSC hospitalizations, accounting for 56.7% (n: 310,580) of cases, followed by infectious diseases and MCNC (**Fig 1**). NCDs also exhibited the highest burden, with in-hospital fatality rates 2 to 4 times higher (3.4%–4.3%) than infectious or maternal-child conditions (0.3%–1.2%) (**S3 Table)**.

**Fig 1.**
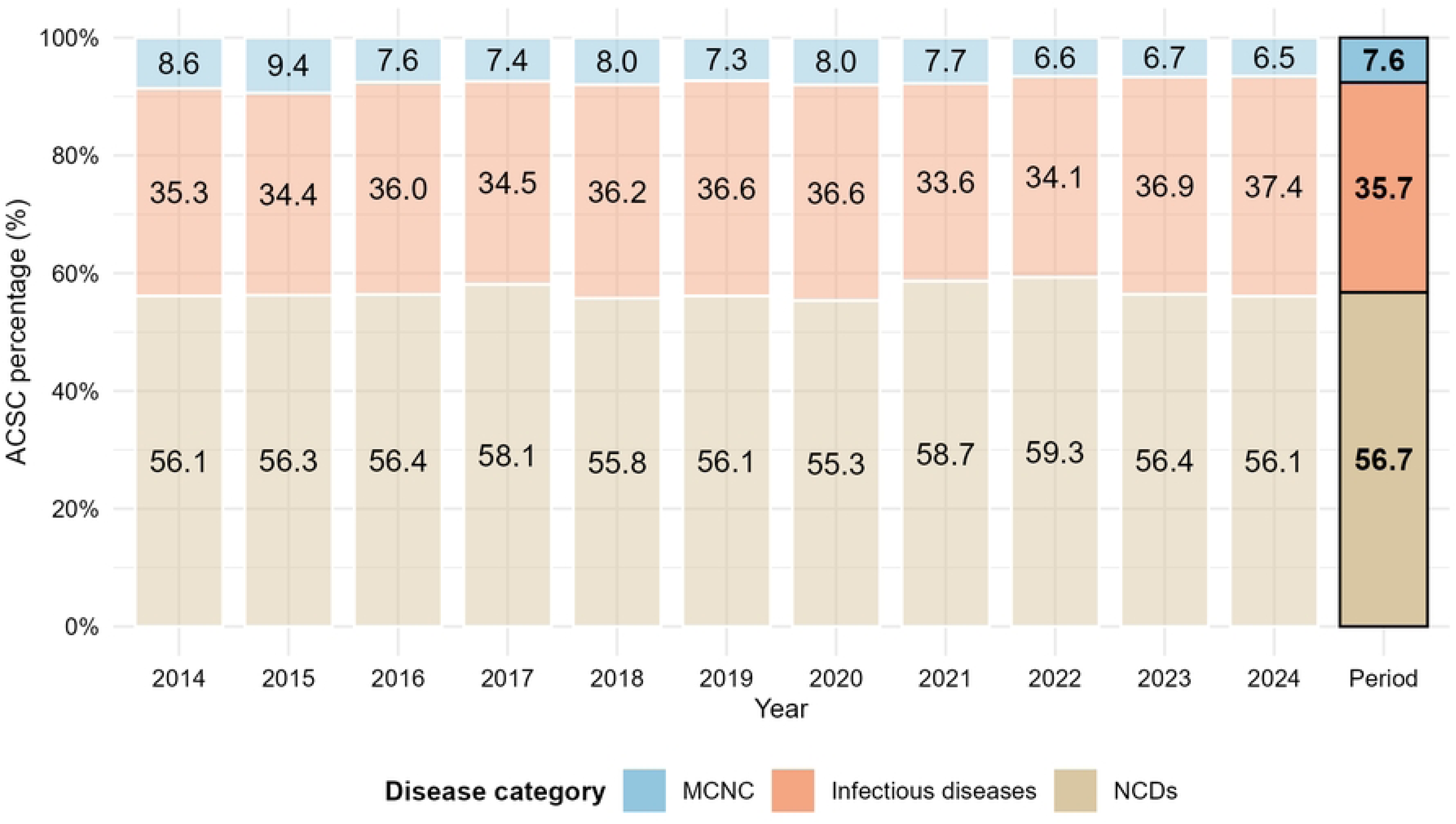
Composition of ACSCs by major disease categories in Honduras, 2014–2024.

The predominant causes of ACSC hospitalizations varied substantially across age groups (**Table 3**). Among children under 5 years, infectious conditions such as gastroenteritis and lower airway diseases accounted for most admissions. In contrast, among adults aged 20 years and older, NCDs, particularly diabetes mellitus and hypertension, represented the largest share of hospitalizations and were associated with higher in-hospital fatality rates. Among individuals aged 80 years and older, cerebrovascular disease exhibited the highest fatality rate (14.7%). A detailed breakdown of ACSC hospitalizations by diagnostic group is provided in **S4 Table**.

**Table 3.**
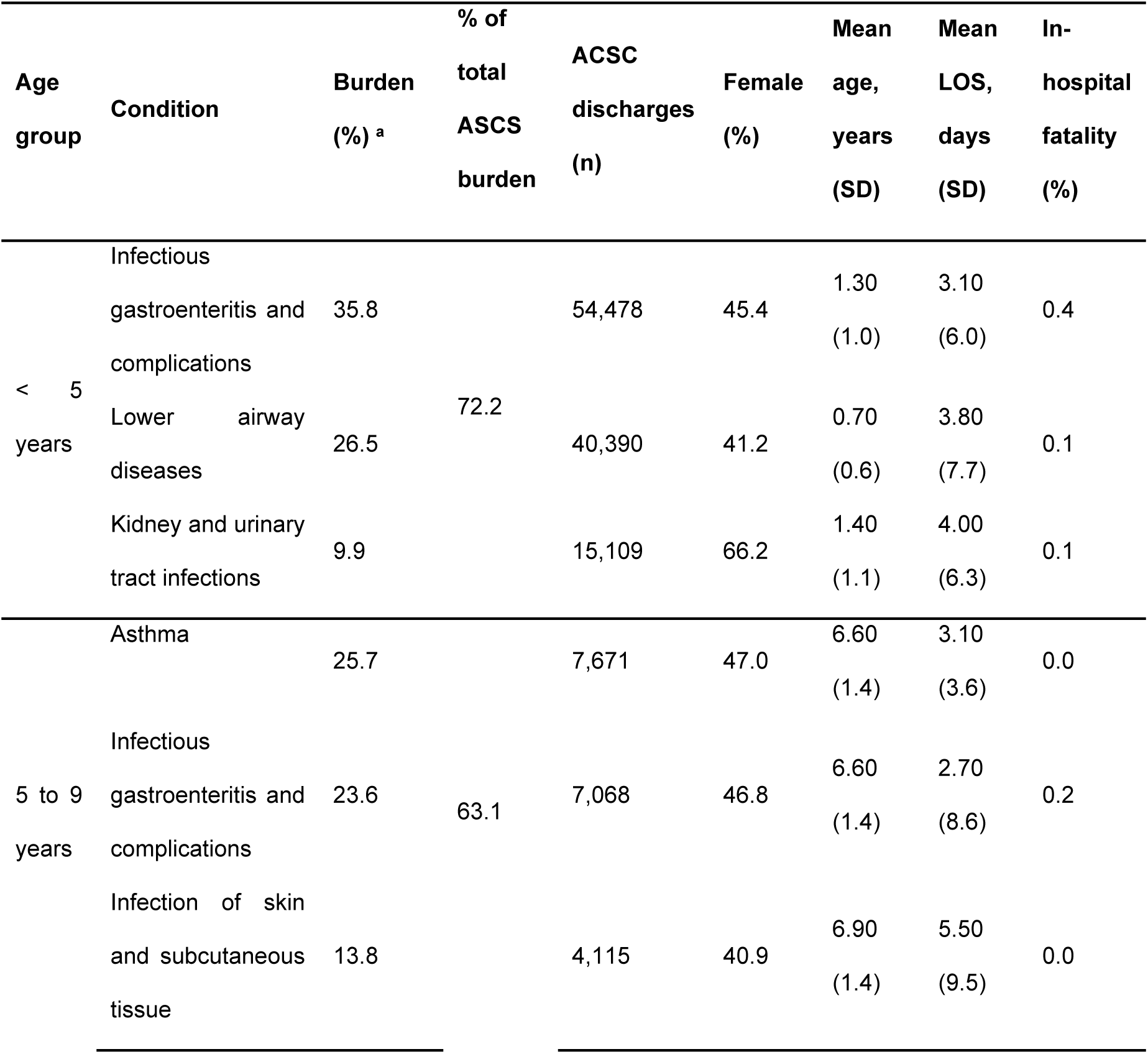
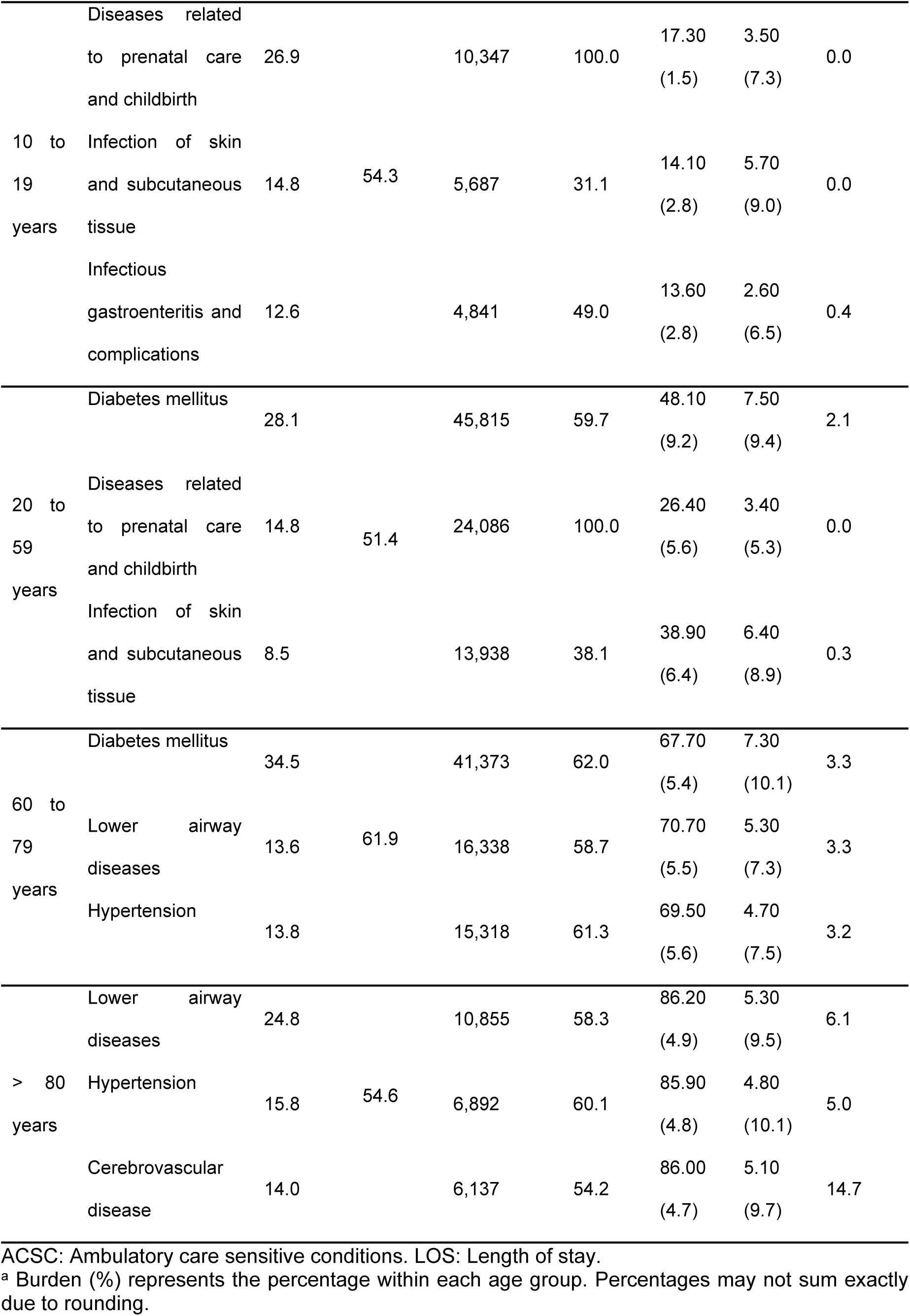
Prevalent ACSC groups and clinical characteristics by age cohort in Honduras, 2014–2024.

Joinpoint regression identified three distinct temporal segments in age-standardized ACSC hospitalization rates over the study period (**Table 4; S1 Fig**). From 2014 to 2018, rates increased but not significantly (APC: 2.7%). This was followed by a sharp and significant decline during the 2018 to 2021 period (APC: -17.8%). Finally, between 2021 and 2024, rates increased significantly again (APC: 15.9%), approximately five times the growth rate of the first period (**Fig 2)**. The overall trend across the full study period was relatively stable (AAPC: -0.4%).

**Table 4.** Trends in age-standardized ACSC hospitalization rates using Joinpoint regression, 2014–2024.

| Outcome | Segment (years) <sup>a</sup> | APC (%) <sup>b</sup> | 95% CI | p-value <sup>c</sup> | Jointpoint year |
| --- | --- | --- | --- | --- | --- |
| ACSC rate (overall) | 2014-2018 | 2.7 | -2.4; 15.2 | 0.295 | 2018 |
|  | 2018-2021 | -17.8 | -30.6; -10.3 | < .001 | 2021 |
|  | 2021-2024 | 15.9 | 4.6; 37.6 | < .001 | -- |
| AAPC (2014-2024) | Total period <sup>d</sup> | -0.4 | -2.4; 2.1 | 0.832 | -- |

AAPC: Average Annual Percent Change. ACSC: Ambulatory care sensitive conditions. APC: Annual Percent Change. CI: Confidence Intervals.
<sup>a</sup> Segments were identified using Joinpoint regression models allowing up to three inflection points.
<sup>b</sup> APC represents the annual change in rates within each segment.
<sup>c</sup> Joinpoints represent statistically significant changes in trend ( $p < 0.05$ ). Rates are age standardized using the 2019 Honduran population as the reference.
<sup>d</sup> AAPC summarizes the overall trend across the full study period.

**Fig 2.**
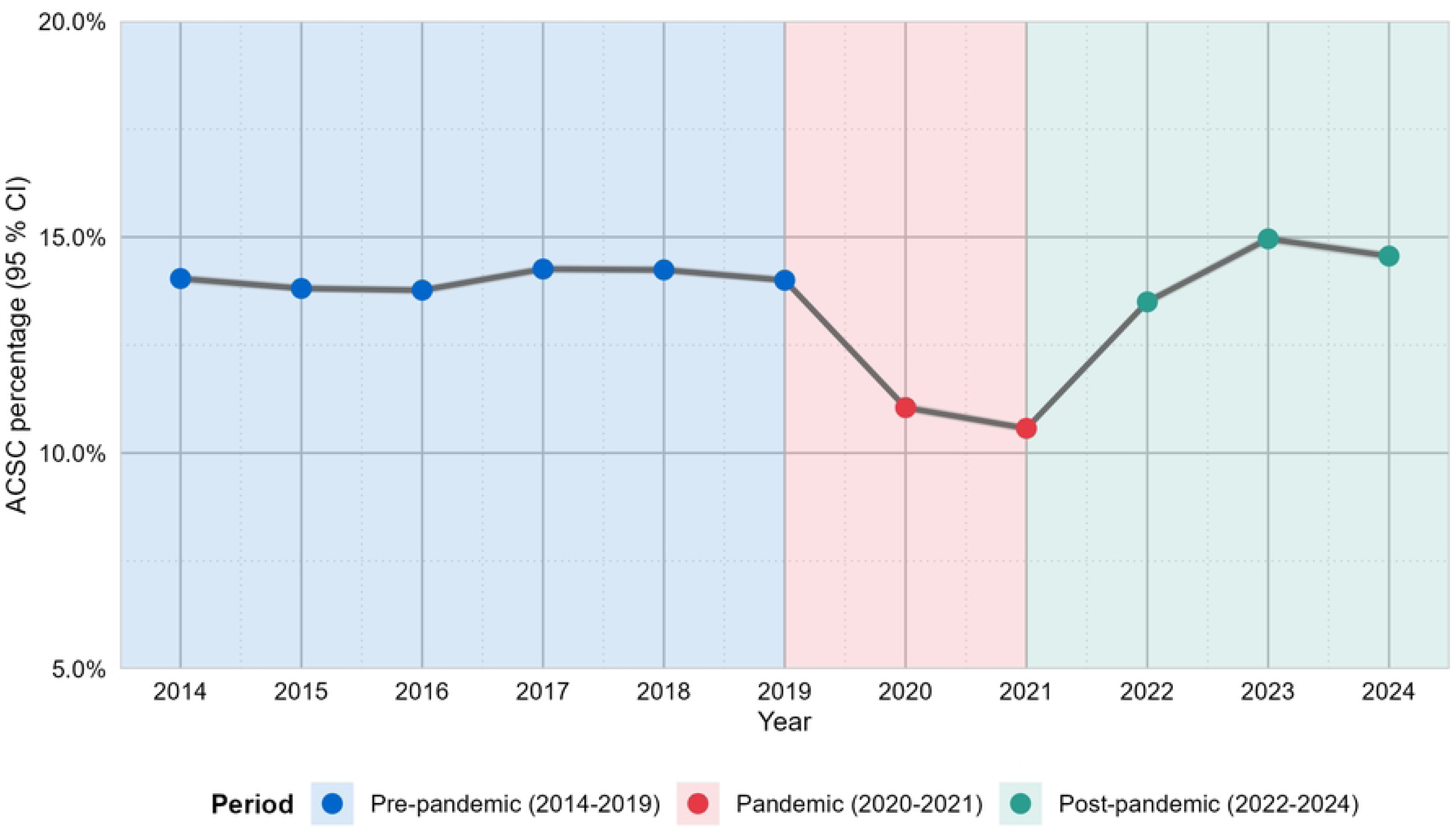
Annual trends in the percentage of Preventable Hospitalizations in Honduras, 2014–2024.

Stratified analyses revealed significant heterogeneity in temporal patterns across population subgroups (**Table 5; S2 Fig**). Among males, age-standardized ACSC hospitalization rates declined sharply and significantly between 2018 and 2020 (APC: -24.6%), followed by an immediate rebound from 2020 to 2024 (APC: 9.6%). In contrast, females experienced a more prolonged decline in ACSC, extending from 2018 to 2021 (APS: -18.5). By disease category, NCD-related ACSC hospitalizations experienced a sharp, significant decline ending in 2020 (APC: -25.3%) and subsequently increased through 2024 (APC: 8.4%). Similar patterns were observed for infectious diseases and MCNC conditions, although the magnitude of changes differed (**Table 5; S3 Fig**).

**Table 5.** Joinpoint regression analysis of age-standardized ACSC hospitalization rates stratified by sex and disease category, 2014–2024.

| Group | Segment (years) <sup>a</sup> | APC (%) <sup>b</sup> | 95% CI | p-value | Joinpoint year |
| --- | --- | --- | --- | --- | --- |
| **Sex** |  |  |  |  |  |
| Male | 2014-2018 | 4.7 | -0.6; 19.7 | 0.071 | 2018 |
|  | 2018-2020 | -24.6 | -33.2; -10. | < 0.001* | 2020 |
|  | 2020-2024 | 9.6 | 2.5; 27.9 | 0.009 | -- |
|  | 2014-2024 | -0.2 | -- | -- | -- |
| Female | 2014-2018 | 2.4 | -2.8; 14.2 | 0.332 | 2018 |
|  | 2018-2021 | -18.5 | -18.5; -31.2 | < 0.001 | 2021 |
|  | 2021-2024 | 15.8 | 4.5; 37.3 | < 0.001 | -- |
|  | 2014-2024 | -0.8 | -2.9; 1.5 | 0.511 | -- |
| **Disease category** |  |  |  |  |  |
| NCDs <sup>d</sup> | 2014-2018 | 3.6 | -1.0; 16.8 | 0.114 | 2018 |
|  | 2018-2020 | -25.4 | -33.3; -11.6 | < .001 | 2020 |
|  | 2020-2024 | 8.4 | 1.6; 25.0 | 0.014 | -- |
|  | 2014-2024 | -1.2 | -- | -- | -- |
| Infectious diseases | 2014-2019 | -3.5 | -7.6; 11.9 | 0.371 | 2019 |
|  | 2019-2021 | -22.8 | -33.6; -10.2 | 0.012 | 2021 |
|  | 2021-2024 | 12.4 | -1.7; 40.1 | 0.077 | -- |
|  | 2014-2024 | 0.8 | -- | -- | -- |
| MCNC <sup>e</sup> | 2014-2019 | 1.5 | -1.9; 9.5 | 0.293 | 2019 |
|  | 2019-2021 | -25.9 | -34.0; -12.3 | < 0.001 | 2021 |
|  | 2021-2024 | 12.4 | -1.7; 40.1 | 0.078 | -- |
|  | 2014-2024 | -1.2 | -- | -- | -- |

ACSC: Ambulatory care sensitive conditions. APC: Annual Percent Change. CI: Confidence Interval. MCNC: Maternal, child, and nutritional conditions. NCDs: Noncommunicable diseases
<sup>a</sup> Segments were identified using Joinpoint regression models.
<sup>b</sup> APC represents the yearly change in rates within each segment.

## Discussion

This study provides the first national assessment of ACSC hospitalizations in Honduras using administrative data from the MoH over an 11-year period. Between 2014 and 2024, ACSCs accounted for 13.6% of all hospitalizations, corresponding to more than half a million avoidable admissions. These findings highlight a substantial and persistent burden of potentially preventable hospitalizations within the Honduran public health system. The magnitude of ACSCs observed in this study underscores important gaps in access to and quality of PHC services. Given that ACSCs are widely used as an indirect indicator of PHC performance, the results suggest that a considerable percentage of hospital admissions may be preventable through timely, continuous, and effective outpatient care. [21–23] In the context of Honduras, where health system fragmentation and barriers to access remain prevalent, [16] these findings are highly relevant for informing strategies to strengthen PHC and improve system efficiency.

### Comparison with regional and global evidence

The sex-age-standardized percentage of ACSCs hospitalizations observed in Honduras (14.9%) is lower than the LAC regional average of 17.4%, as reported in a multicountry study covering 2015 to 2019. [6] However, when compared with high-income settings, the burden in Honduras remains substantially higher. Studies from Europe have reported ACSC percentages ranging from 4.5% in countries such as Spain and the United Kingdom to approximately 11.0% in countries such as Portugal. [24–29] In Oceania, Australia has documented a percentage of approximately 6.0%, [30] while estimates in Asia range from 4.2% in South Korea to 8.4% in Japan [31–33].

These differences may reflect structural disparities in health system organization, particularly in the strength and accessibility of PHC services. High-income countries typically have more consolidated primary care systems with better care coordination, continuity, and preventive services, which contribute to lower rates of avoidable hospitalizations. [24–26,34] In contrast, the higher burden observed in Honduras and other countries in the region may be driven by persistent barriers to timely access, limited continuity of care, and resource constraints within PHC systems.

### Trends before, during, and after the COVID-19 pandemic

The temporal analysis revealed three distinct phases in ACSC hospitalization trends, with a marked disruption during the COVID-19 pandemic. Between 2014 and 2018, ACSC hospitalization rates increased steadily, followed by a sharp decline between 2018 and 2021, and a subsequent recovery during the post-pandemic period (2021 and 2024).

The identification of 2018 as the initial joinpoint for the decline may be attributed to both statistical and epidemiological factors. Mathematically, the joinpoint regression algorithm experiences a “leverage effect” when faced with a massive, non-linear drop, such as the one observed in 2020. [35] To minimize the total sum of squared errors across the segment, the model ”anticipates” the trajectory shift by placing the node at the 2018 mark. From a public health perspective, this shift also coincides with the 2019 Dengue emergency in Honduras—the most severe outbreak in the country’s recent history. [36] This crisis likely saturated the healthcare system and prioritized acute epidemic management, potentially displacing elective care and disrupting the baseline stability of other ACSC admissions just before the total systemic shock triggered by COVID-19.

The decline observed during the pandemic is consistent with international evidence documenting substantial reductions in hospital utilization, including avoidable hospitalizations, during this period. [21] A study conducted in England found that the pandemic led to significant changes in ACSC distribution across regions, influenced by socioeconomic and demographic factors. [37] Similarly, evidence from China suggests that the pandemic altered hospital service utilization patterns and revealed associations between ACSC rates and socioeconomic characteristics. [32] Multiple factors may explain this pattern, including reduced healthcare-seeking behavior due to fear of infection, mobility restrictions, and the reallocation of health system resources toward the COVID-19 response. [37,38]

The post-pandemic rebound observed in Honduras, particularly the accelerated increase in ACSC rates after 2021, may reflect delayed care, worsening of chronic conditions, and accumulated unmet healthcare needs during the pandemic period. [32] Supporting this interpretation, a study in the United States found that older adults had a substantially increased risk of developing ACSCs following COVID-19 infection, both in the short and long term. [39] These dynamics highlight the lasting impact of the pandemic on health system performance and population health.

### Clinical and demographic patterns

The clinical profile of ACSCs in Honduras is characterized by a predominance of NCDs, which accounted for 56.7% of all cases, followed by infectious diseases and MCNC. This distribution reflects the ongoing epidemiological transition in the country, where chronic conditions increasingly coexist with infectious diseases. The most frequent diagnostic groups, including diabetes mellitus, infectious gastroenteritis, lower airway diseases, urinary tract infections, and skin infections, are consistent with patterns reported in other LAC countries. For example, regional evidence indicates that diabetes mellitus represents the leading cause of ACSC, followed by urinary tract infections, gastroenteritis, and skin infections, suggesting a broadly similar epidemiological profile across the region [6].

Important differences emerged across age groups. ACSC were disproportionately concentrated on the extremes of the life course, particularly among children under five years and older adults. In younger populations, infectious conditions predominated, reflecting vulnerabilities related to environmental exposures, nutrition, and access to preventive care. In contrast, among adults and older populations, NCDs such as diabetes and hypertension were the primary drivers of ACSC hospitalizations and were associated with higher fatality rates. [6,40] These findings underscore the need for age-specific strategies within PHC to address both communicable and noncommunicable disease burdens.

### In-hospital fatality and length of stay

ACSC hospitalizations in Honduras were associated with a higher in-hospital fatality rate compared with non-ACSC hospitalizations, as well as longer LOS. These findings suggest that, although theoretically preventable, ACSC cases often present at more advanced stages or with greater clinical severity when hospitalization occurs. The observed fatality rates are consistent with findings from other countries in the LAC region. For example, a study in Ecuador reported an in-hospital fatality rate of approximately 2% among ACSC hospitalizations, [41] while evidence from Colombia indicates a range between 0.5% and 4.6% depending on region and health insurance scheme. [42] Similarly, the average LOS observed in this study falls within the range reported in multicountry analyses in the Americas (4.6 to 6.8 days) [43]. Interestingly, unlike findings from China that documented an increase in hospital LOS during the pandemic followed by a return to baseline levels, [32] the Honduran data showed a slight but consistent decline in length of stay over time. This pattern may reflect changes in hospital management practices or constraints on bed availability, although further research is needed to better understand these dynamics.

### Policy implications

The findings of this study have several important implications for health policy in Honduras. First, the substantial burden of ACSCs underscores the need to strengthen PHC capacity, particularly for managing chronic diseases such as diabetes and hypertension. Expanding access to continuous care, improving early detection, and ensuring treatment adherence could reduce preventable hospitalizations and improve health outcomes. [21,22,44,45] Second, the concentration of ACSCs among young children and older adults highlights priority populations for targeted interventions. Strengthening preventive services, vaccination programs, and integrated care models for these groups could yield significant gains in reducing ACSCs burden. Third, the marked disruption and subsequent rebound associated with the COVID-19 pandemic emphasize the need to build more resilient health systems capable of maintaining essential services during crises. This includes strengthening service integration, improving surveillance systems, and ensuring continuity of care during emergencies.

### Strengths, limitations, and future research directions

This study has several important strengths. It represents the first national analysis of ACSC hospitalizations in Honduras and is based on a large administrative dataset covering more than four million hospitalizations over an 11-year period. The use of a standardized ACSC classification based on ICD-10 codes enhances comparability with other studies in the region and globally.

However, several limitations should be considered. First, the analysis relies on administrative data, which may be subject to coding errors and does not capture the full clinical complexity of patients. Second, the data lack information on individual-level socioeconomic and behavioral factors, limiting the ability to explore underlying determinants of ACSC hospitalizations. Third, the study includes only hospitalizations within the MoH network and does not capture cases from the Honduran Social Security Institute or private sector, which may limit generalizability. Finally, ACSC hospitalizations should be interpreted as an indirect proxy of PHC performance rather than a direct measure of system functioning. Hospitalization decisions are influenced by multiple factors, including clinical judgment, hospital capacity, and patient characteristics. Therefore, the findings should be interpreted cautiously and complemented with additional sources of evidence.

The findings of this study highlight several priorities for future research. First, subnational analyses are needed to better understand geographic variation in ACSC rates and identify areas with the greatest need for intervention. Second, studies incorporating socioeconomic and health system variables could help identify key determinants of ACSC hospitalizations and inform targeted policies. Third, economic evaluations are needed to quantify the financial burden of ACSCs on the health system and society, including both direct costs and indirect productivity losses.

### Conclusions

ACSCs account for a substantial percentage of hospitalizations within the Honduran public health system. Although ACSC rates declined during the COVID-19 pandemic, the subsequent rebound highlights the sensitivity of this indicator to disruptions in health system functioning. The predominance of NCDs and the concentration of cases among young children and older adults underscore the need for targeted and integrated PHC interventions. Strengthening primary care services, improving chronic disease management, and enhancing health system resilience are essential steps to reduce preventable hospitalizations and advance toward more efficient and equitable health care in Honduras.

### Authors’ notes

In line with calls to advance language equity in health research, [46] we provide a full Spanish translation of this review as supplementary material to support equitable evidence access and policy uptake beyond English-speaking audiences (**S1 File**).

## Acknowledgments section

This study was made possible by the support and collaboration of various actors within the Honduran Ministry of Health. We thank them all for their time, support, and contribution.

## Data availability

The data that support the findings of this study consist of national hospital discharge records from the Honduran Ministry of Health (MoH). These data were obtained under a specific data-sharing agreement with the MoH, and the authors had permission to use the data for the purposes of this study. The data are not publicly available and cannot be shared by the authors due to institutional restrictions. Researchers interested in accessing the data may submit a request directly to the Health Statistics Department of the Honduran MoH, subject to their approval and applicable regulations, through the following contact portal: https://www.salud.gob.hn/sshome/index.php/contactanos

## Supporting information

**S1 Table.** Definition of Condition Groups and Specific ICD-10 Codes for ACSC.

**S2 Table.** Annual Population Denominators and Estimates for ACSC Rate Calculations in Honduras, 2014–2024.

**S3 Table.** Annual National Characteristics and Demographic Profile of ACSC Hospitalizations in Honduras, 2014–2024.

**S4 Table.** Clinical Characteristics and Outcomes of ACSCs in Honduras by Diagnostic Group, 2014–2024.

**S1 Fig.** Joinpoint Trend Analysis of Age-Standardized ACSC Hospitalizations Rates in Honduras, 2014-2024

**S2 Fig.** Joinpoint Trend Analysis of Age-Standardized ACSC Hospitalization Rates stratified by Sex in Honduras, 2014–2024.

**S3 Fig.** Joinpoint Trend Analysis of Age-Standardized ACSC Hospitalization Rates stratified by Disease Category in Honduras, 2014–2024.

**S1 File.** Full Spanish translation of the article.

